# Predictors of Vaginal Birth at the Time of Admission in a Contemporary Cohort of Term Nulliparas

**DOI:** 10.1101/2025.04.25.25326456

**Authors:** Nadine Sunji, Amy Y. Pan, Liyun Zhang, Margaret H. Bublitz, Nina Ayala, Abbey Kruper, Anna Palatnik

## Abstract

**Background:** Designing a model to accurately predict vaginal birth in nulliparas may play a role in reducing cesarean delivery rates. This study aimed to identify predictors of vaginal birth at the time of admission in a contemporary and diverse cohort of nulliparous patients in the United States, and to assess the accuracy of the developed model.

**Methods:** This was a retrospective cohort study from the Nulliparous Pregnancy Outcomes Study: Monitoring Mothers-to-be. The present analysis included nulliparous patients with live, singleton, non-anomalous, term gestations, and no contraindications to vaginal birth. Candidate predictors were selected based on clinical knowledge and literature review. Bivariable analyses and multivariable logistic regression were used to establish factors independently associated with vaginal birth. A receiver operating characteristic curve was generated, and the area under the curve was calculated to estimate the predictive capacity of the final model.

**Results:** Among the 6,043 individuals included, 5,058 (83.7%) delivered vaginally and 985 (16.3%) delivered via cesarean. Independent predictors of vaginal birth included maternal age, maternal height, body mass index at delivery, gestational age at delivery, group B Streptococcus status, chorioamnionitis, gestational hypertension/preeclampsia, diabetes, cervical dilation, cervical effacement, and fetal sex. The final predictive model achieved an area under the receiver operating characteristic curve of 0.79 (95% confidence interval, 0.77–0.81).

**Conclusion:** Several factors, most available prior to labor, were identified to be independently associated with vaginal birth at the time of admission among term nulliparous patients. Combining these independent predictors resulted in a clinical prediction model with fair predictive capability.

## Introduction

Nulliparous patients currently exhibit increasing cesarean delivery rates.^1^ In 2021, the primary cesarean delivery rate in the United States was 22.3 percent with nulliparous, term, singleton, and cephalic births comprising 26.3 percent.^2^ Previous studies’ report multiple reasons for the rise in cesarean rates, including maternal request,^3–5^ physician’s preference or financial incentive,^6^ sociocultural influence,^7, 8^ and fear of malpractice liability.^9–12^ Increased maternal age,^13–15^ obesity,^15^ induction of labor,^13, 16^ diabetes,^13, 15^ hypertensive disorders of pregnancy,^13, 15^ and infertility treatment^14^ have also previously been noted to contribute to the increased rates of nulliparous cesarean delivery.

Individuals who deliver via cesarean face an increased risk of not only short- and long-term postpartum complications but also repeat cesarean deliveries in future pregnancies. These individuals exhibit higher rates of infection,^17^ hemorrhage,^18, 19^ slower recovery time,^20^ and hospital readmission,^21^ and are more likely to suffer complications such as placenta previa or uterine rupture in future pregnancies.^22^ Moreover, babies born via cesarean delivery face added risk of respiratory complications, altered immunity, and prolonged stays in neonatal intensive care units compared with those delivered vaginally.^23^ Cesarean deliveries also increase healthcare expenditures with labor and delivery costing at least 1.5 times the cost of vaginal births and postpartum care at least four times.^24^ Compared with cesarean delivery, vaginal birth lowers the risk of maternal and neonatal morbidity and mortality, enhances the childbirth experience, and reduces healthcare costs. Moreover, given that mode of delivery greatly affects the course of future pregnancies, determining factors that influence the success of vaginal birth is crucial and may aid in reducing the overall cesarean delivery rates. Nulliparas’ considerable contribution to cesarean delivery rates highlights the need for the identification of strategies to reduce these rates.

Several investigators have previously described prediction models for vaginal or cesarean delivery in specific patient populations, such as patients with type 1 diabetes mellitus,^25^ patients undergoing induced labor,^15, 26–29^ and patients with preterm pregnancies.^30^ Models predicting vaginal birth in nulliparous individuals have also been developed and reported significant predictors such as maternal age,^28, 31^ race,^28^ height,^28^ gestational age at delivery,^27, 28, 31^ cervical exam,^28, 31^ hypertension,^28^ and diabetes.^28^ However, prior findings were often limited to small cohorts from single institutions and/or to certain patient populations. Therefore, this study aimed to identify independent predictors of vaginal birth at the time of admission in a contemporary and diverse cohort of term nulliparous patients in the United States, and to assess the accuracy of the prediction model developed.

## Methods

This was a secondary analysis of the Nulliparous Pregnancy Outcomes Study: Monitoring Mothers-to-be (nuMoM2b). The nuMoM2b prospective cohort study collected demographic, psychosocial, dietary, physiologic, and pregnancy outcome data from 9,289 nulliparous (no previous pregnancy lasting ≥20 weeks’ gestation) people from eight clinical sites in the United States between 2010 and 2014 to investigate adverse pregnancy outcomes.^32^ Nulliparous individuals with a viable singleton pregnancy were recruited during the first trimester of pregnancy and were followed at four different timepoints during pregnancy until delivery. Data was collected through interviews, self-administered questionnaires, clinical measurements, ultrasound imaging, and review of medical records. Additional information about the nuMoM2b study’s design and methodology can be found in previous publications.^33^ The Institutional Review Board at the Medical College of Wisconsin and the Eunice Kennedy Shriver National Institute of Child Health and Human Development (NICHD) approved this secondary analysis.

The present analysis included nulliparous participants with live, non-anomalous, singleton gestations at term (≥37 weeks’ gestation at birth). Participants with planned cesarean delivery and contraindications to vaginal birth (namely placenta accreta, placenta previa, vasa previa, umbilical cord prolapse, fetal malpresentation, herpes simplex virus, human immunodeficiency virus, and prior myomectomy) were excluded. Individuals with eclampsia, hemolysis, elevated liver enzyme, low platelet (HELLP) syndrome, cardiac disease, or coronary artery disease were also excluded because of high rates of cesarean delivery associated with these diagnoses.

The primary outcome of interest was the predictive model of vaginal birth. Maternal sociodemographic data, labor and delivery characteristics, complications present during pregnancy, cervical characteristics, and fetal characteristics were abstracted from the dataset and were compared between individuals who delivered vaginally and those who delivered via cesarean. Candidate predictors were selected based on clinical knowledge and literature review. More specifically, only factors typically available at labor were selected as potential predictors of vaginal birth as they can inform clinical decisions prior to the attempt of a vaginal birth.

To identify factors associated with vaginal birth, a bivariable analysis was performed using chi-square test and Mann-Whitney *U* test as appropriate. All variables found to be significantly associated with vaginal birth (*p*-value<.05) were included in the multivariable logistic regression model. A backward selection was conducted to identify predictors significantly associated with vaginal birth. Restricted cubic spline function was used to model continuous variables when a nonlinear association was detected. Using the final regression model, a receiver operating characteristic (ROC) curve was then generated, and the area under the curve (AUC) was calculated to determine the predictive capacity of the final model. Internal validation was conducted using 1,000 bootstrap resamples, and the optimism-corrected C-statistic (calculated as the naïve C-statistic minus the estimated optimism) was reported. A nomogram was constructed based on the final prediction model. An AUC of at least 0.80 was considered sufficient for clinical applicability.^34^ All analyses were performed with the software SAS version 9.4 (SAS Institute Inc., Cary, NC) and R version 4.4.1 (R Core Team, Vienna, Austria). All tests were two-tailed, and a *p*-value below 0.05 defined statistical significance.

## Results

Of 9,289 nulliparous participants from the nuMoM2b dataset, 6,043 met inclusion criteria and remained in this secondary analysis. Of these, 5,058 (83.7%) participants delivered vaginally, 1,978 (39.1%) of whom underwent induction of labor, and 985 (16.3%) delivered via cesarean, 560 (56.9%) of whom were induced. Demographic and clinical characteristics of the included pregnancies stratified by mode of delivery are presented in Table 1.

**Table 1.**
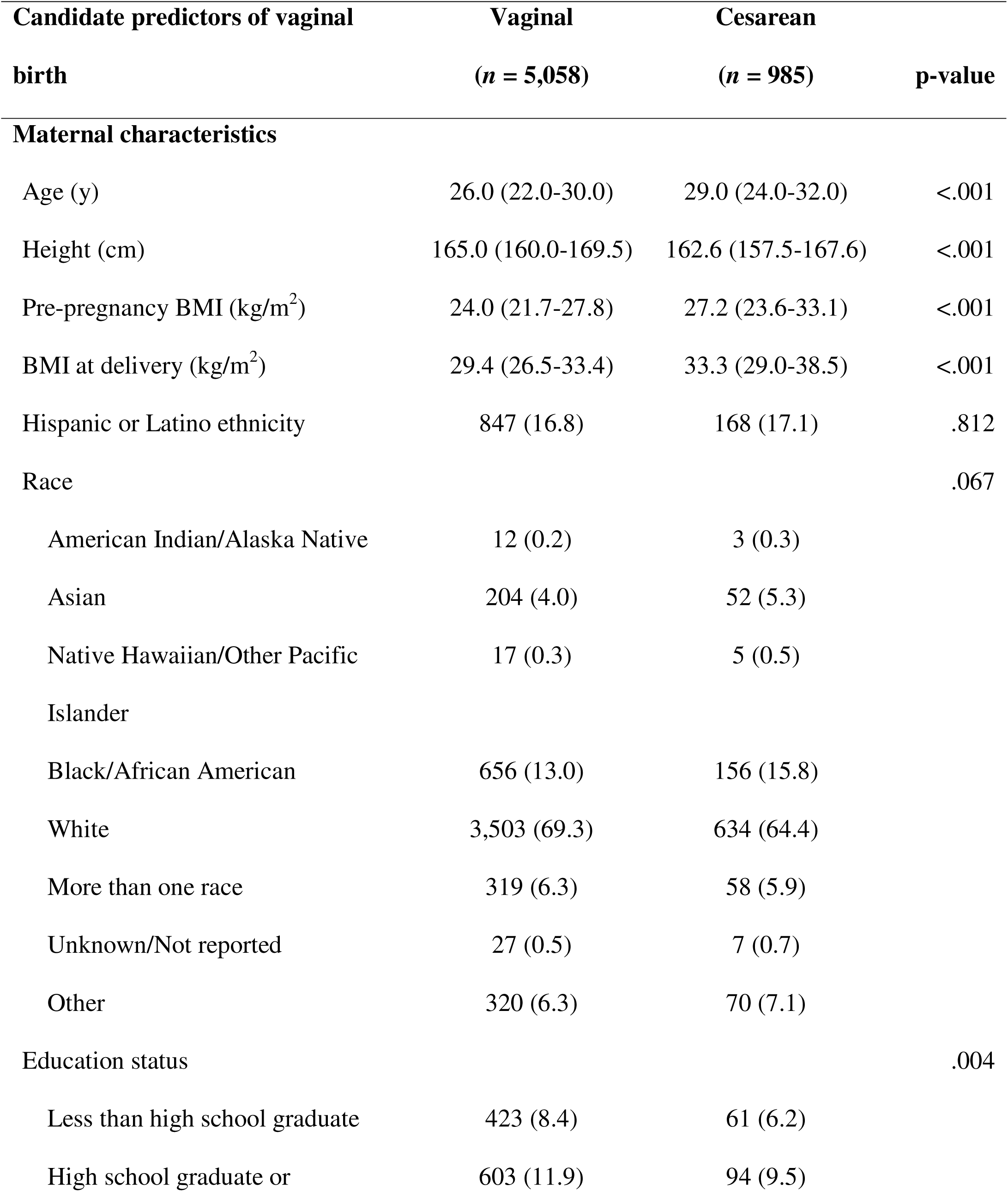
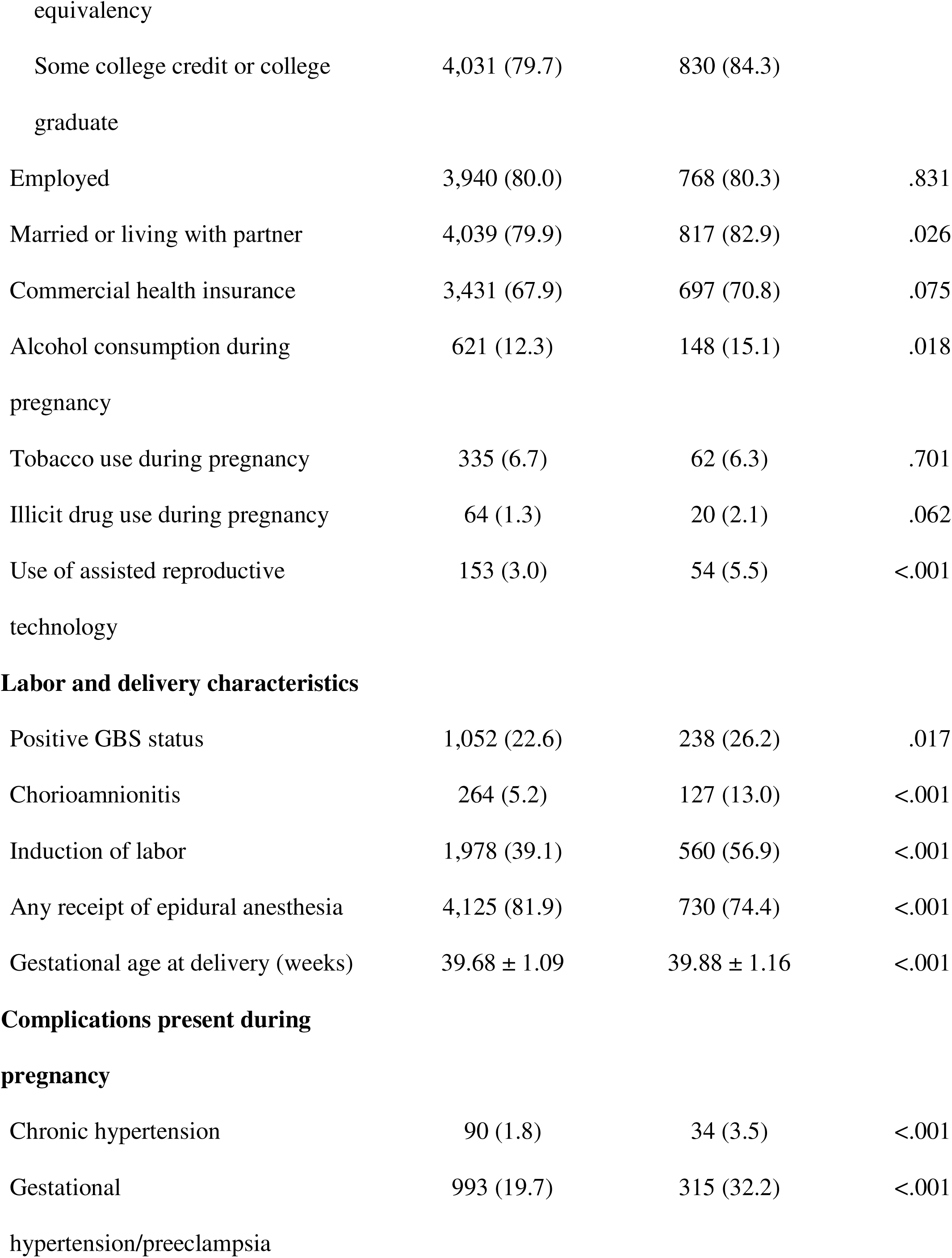
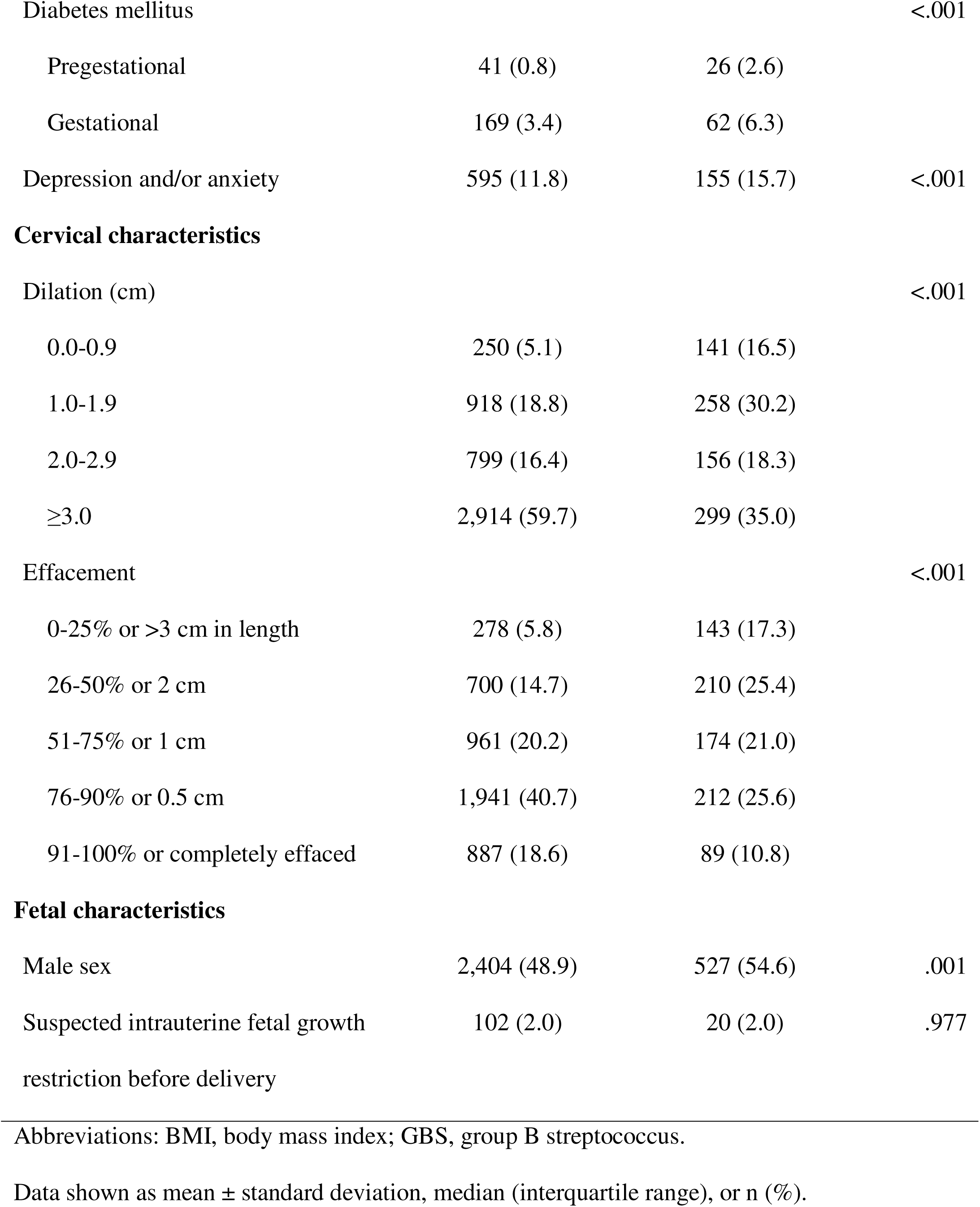
Demographic and clinical characteristics of pregnancies stratified by mode of delivery.

In bivariable analysis, younger maternal age, greater maternal height, lower BMI before pregnancy and at delivery, and lower education level were all associated with higher rates of vaginal birth (all *p*<.010). Individuals who delivered vaginally were also less likely to be married or living with a partner (*p*=.026) and to have consumed alcohol during the current pregnancy (*p*=.018). Individuals who delivered vaginally were more likely to receive epidural anesthesia, to deliver at an earlier gestational age, to have greater cervical dilation and effacement at the start of labor, and to have a female fetus (all *p*≤.001). The use of assisted reproductive technology, induction of labor, positive group B Streptococcus (GBS) status (*p*=.017), and chorioamnionitis were less common in individuals who delivered vaginally (all *p*<.001). Participants who delivered vaginally were also less likely to have complications during pregnancy, namely chronic hypertension, gestational hypertension/preeclampsia, diabetes mellitus (pregestational and gestational), and depression and/or anxiety (all *p*<.001).

After controlling for confounding variables in the multivariable analysis, the following eleven factors remained independently associated with vaginal birth in the final model: maternal age, maternal height, BMI at delivery, gestational age at delivery, GBS status, chorioamnionitis, gestational hypertension/preeclampsia, diabetes, cervical dilation, cervical effacement, and fetal sex (Table 2). The odds of vaginal birth decreased with every 1-year increase in maternal age (aOR = 0.88, 95% confidence interval [CI], 0.78–1.00 with age in 20s; aOR = 0.94, 95% CI, 0.88–0.99 with age in 30s; and aOR = 1.15, 95% CI, 0.82–1.62 with age in 40s) and increased with every 5-cm increase in maternal height (aOR = 1.39, 95% CI, 1.10–1.75 with maternal height in 150s cm; aOR = 1.54, 95% CI, 1.24–1.92 with maternal height in 160s cm; and aOR = 1.38, 95% CI, 1.09–1.74 with maternal height in 170s cm). The odds of vaginal birth decreased with every 1-unit increase in BMI at delivery at BMI levels 25 (aOR = 0.88, 95% CI, 0.78–0.99), 30 (aOR = 0.90, 95% CI, 0.86–0.94), and 35 (aOR = 0.92, 95% CI, 0.89–0.95) but was unaffected at BMI levels 40 (aOR = 1.00, 95% CI, 0.97–1.02) and 45 (aOR = 1.01, 95% CI, 0.97–1.04). The odds of vaginal birth also decreased with every 1-week increase in gestational age at delivery (aOR = 0.82, 95% CI, 0.57–1.17 with gestational age at 37; aOR = 0.85, 95% CI, 0.69–1.04 with gestational age at 38; aOR = 0.88, 95% CI, 0.68–1.15 with gestational age at 39; aOR = 0.70, 95% CI, 0.56–0.88 with gestational age at 40; aOR = 0.68, 95% CI, 0.46–0.99 with gestational age at 41; and aOR = 0.68, 95% CI, 0.46–1.00 with gestational age at 42). Positive GBS status (aOR = 0.80, 95% CI, 0.66–0.98), chorioamnionitis (aOR = 0.38, 95% CI, 0.29– 0.51), and male fetal sex (aOR = 0.70, 95% CI, 0.59–0.84) were associated with lower odds of vaginal birth. Greater cervical dilation of 1.0-1.9 cm (aOR = 1.60, 95% CI, 1.17–2.20), 2.0-2.9 cm (aOR = 1.62, 95% CI, 1.14–2.31), and ≥3.0 cm (aOR = 2.49, 95% CI, 1.76–3.52) were associated with higher odds of vaginal birth compared with dilation of <1.0 cm. Greater cervical effacement of 26-50% or 2 cm in length (aOR = 1.41, 95% CI, 1.04–1.92), 51-75% or 1 cm (aOR = 1.92, 95% CI, 1.38–2.68), 76-90% or 0.5 cm (aOR = 2.29, 95% CI, 1.63–3.23), and 91-100% or complete effacement (aOR = 2.10, 95% CI, 1.40–3.13) were associated with higher odds of vaginal birth compared with 0-25% effacement or >3 cm in length. Pregestational diabetes and gestational hypertension/preeclampsia were both associated with lower odds of vaginal birth (aOR = 0.30, 95% CI, 0.16–0.58 and aOR = 0.57, 95% CI, 0.47–0.69, respectively).

**Table 2.**
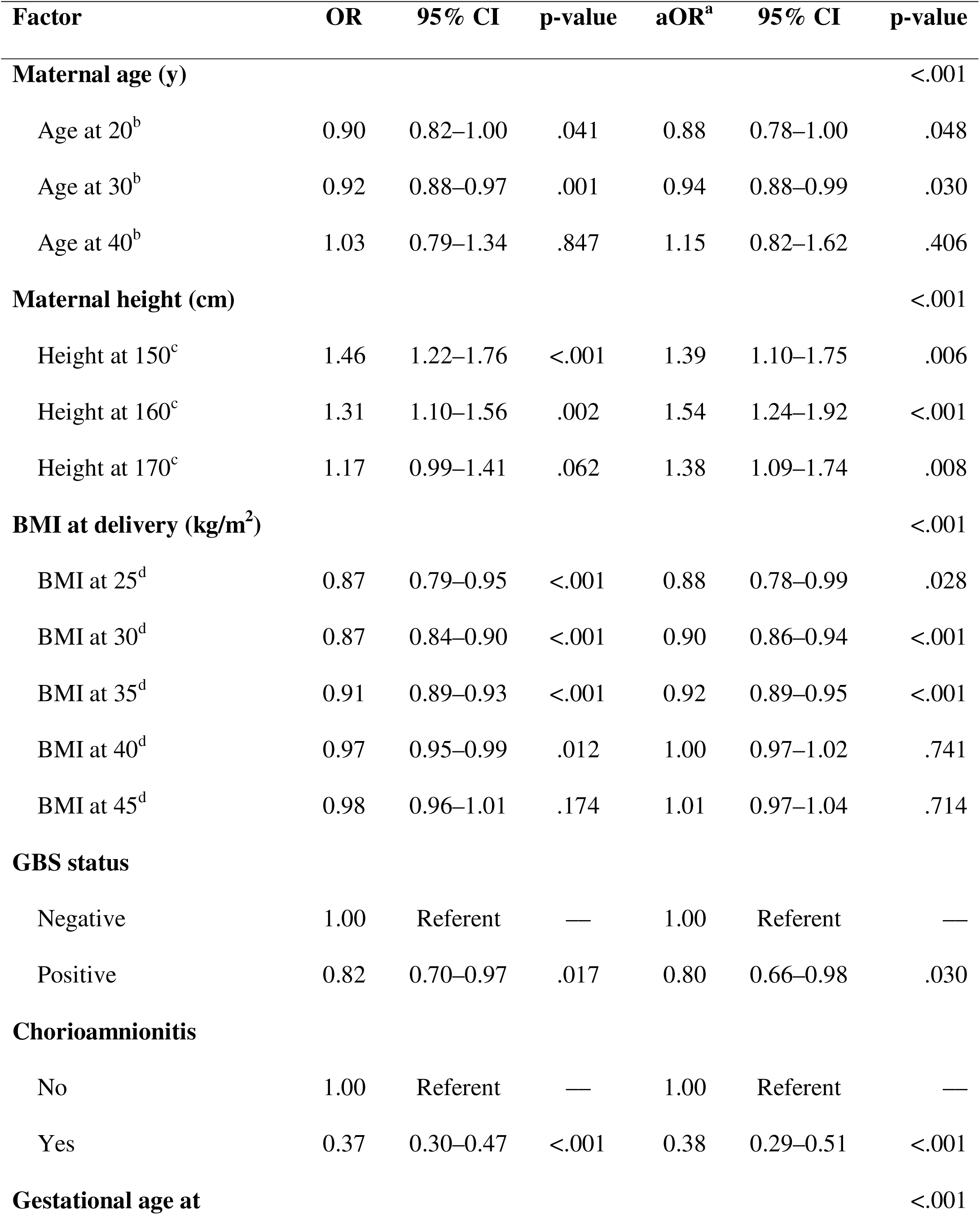
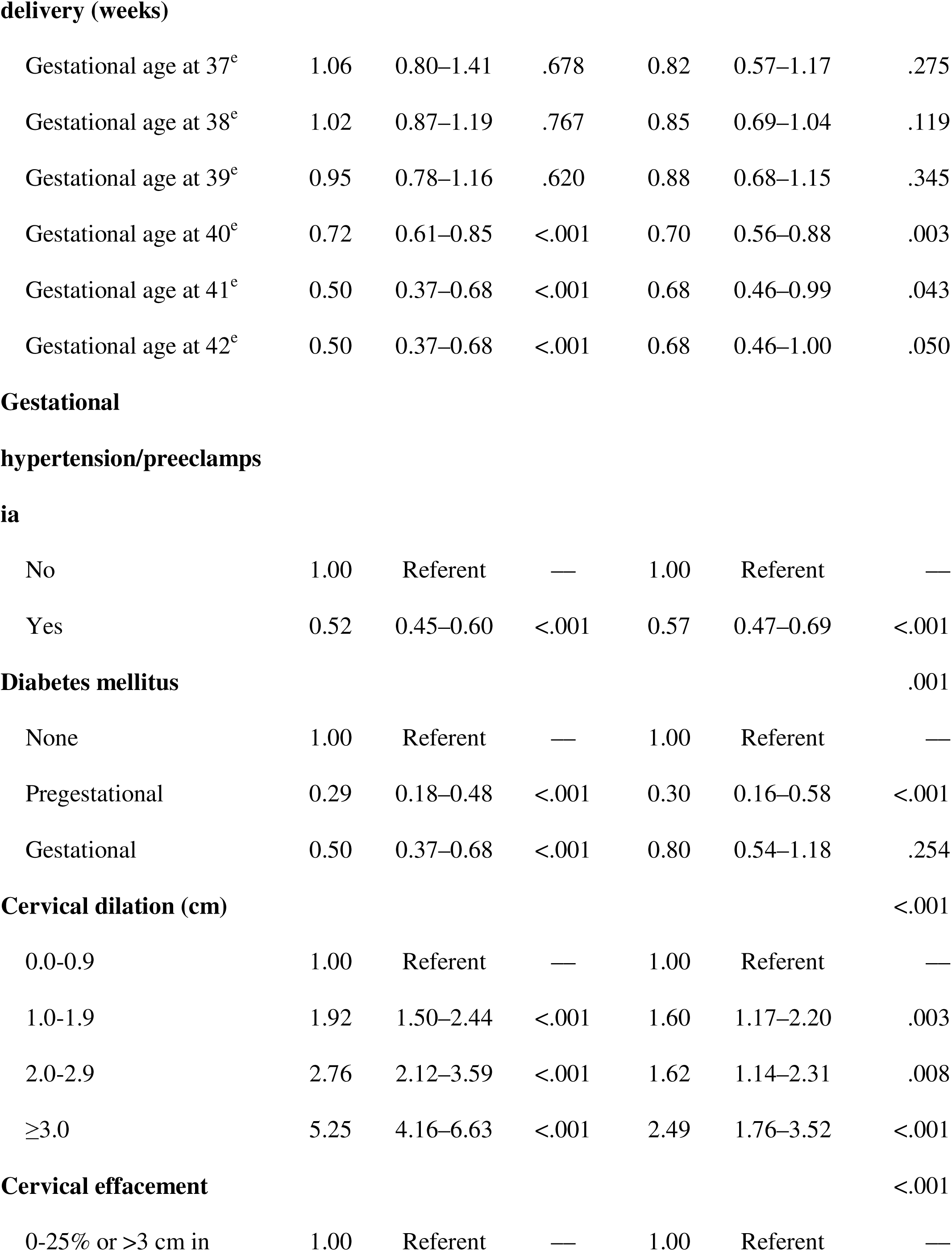
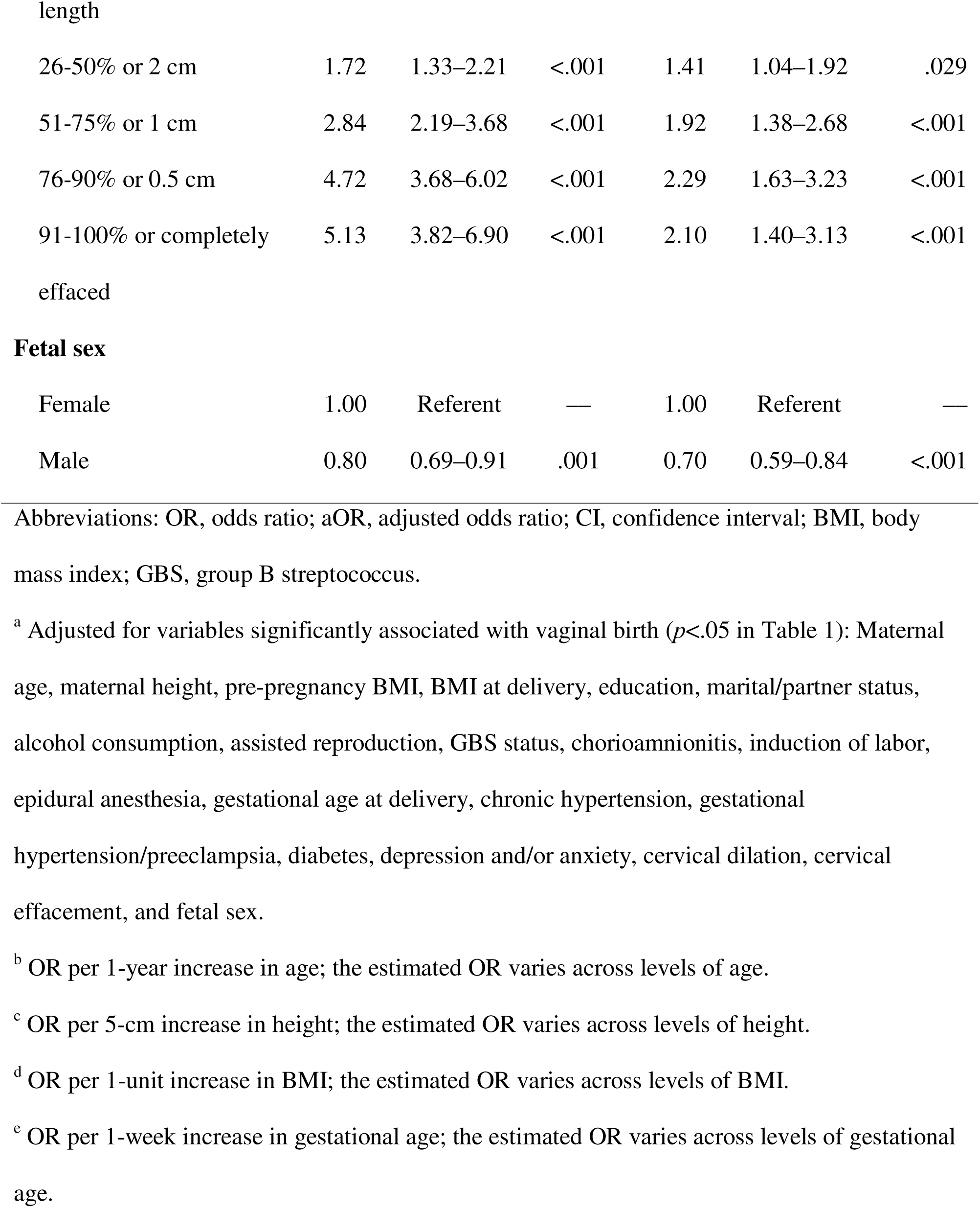
Predictors of vaginal birth in term nulliparas (final model)

Figure 1 demonstrates the ROC curve for final predictive model of vaginal birth in term nulliparas. The AUC was 0.79 (95% CI, 0.77–0.81), and the optimism-corrected C-statistic derived from 1,000 bootstrap resamples was 0.78. Figure 2 shows a predictive graphic nomogram derived from the final model.

**Figure 1.**
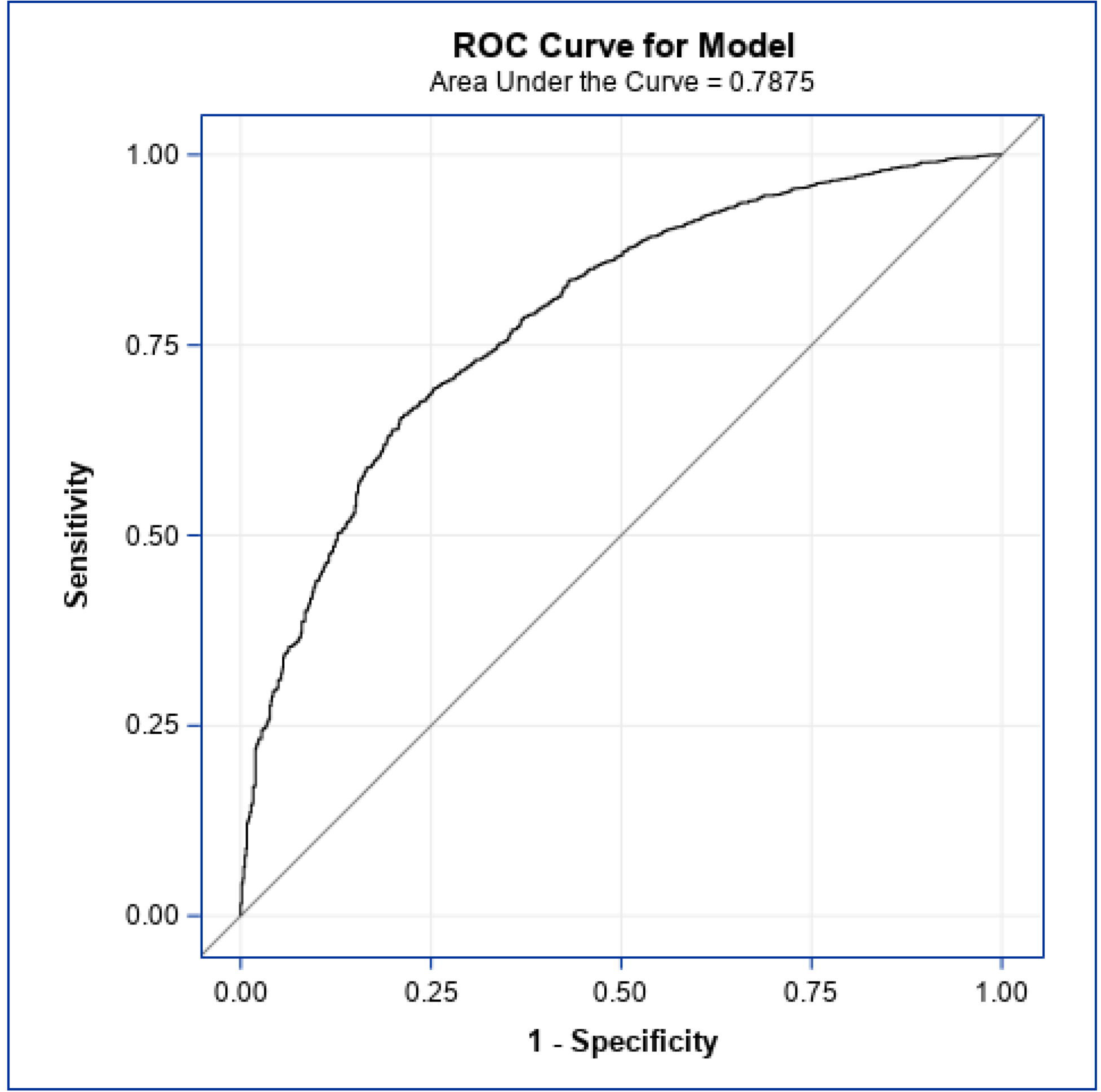
Receiver operating characteristic curve for prediction of vaginal birth in term nulliparas. Area under receiver operating characteristic (ROC) curve = 0.788 (95% confidence interval, 0.770–0.805).

**Figure 2.**
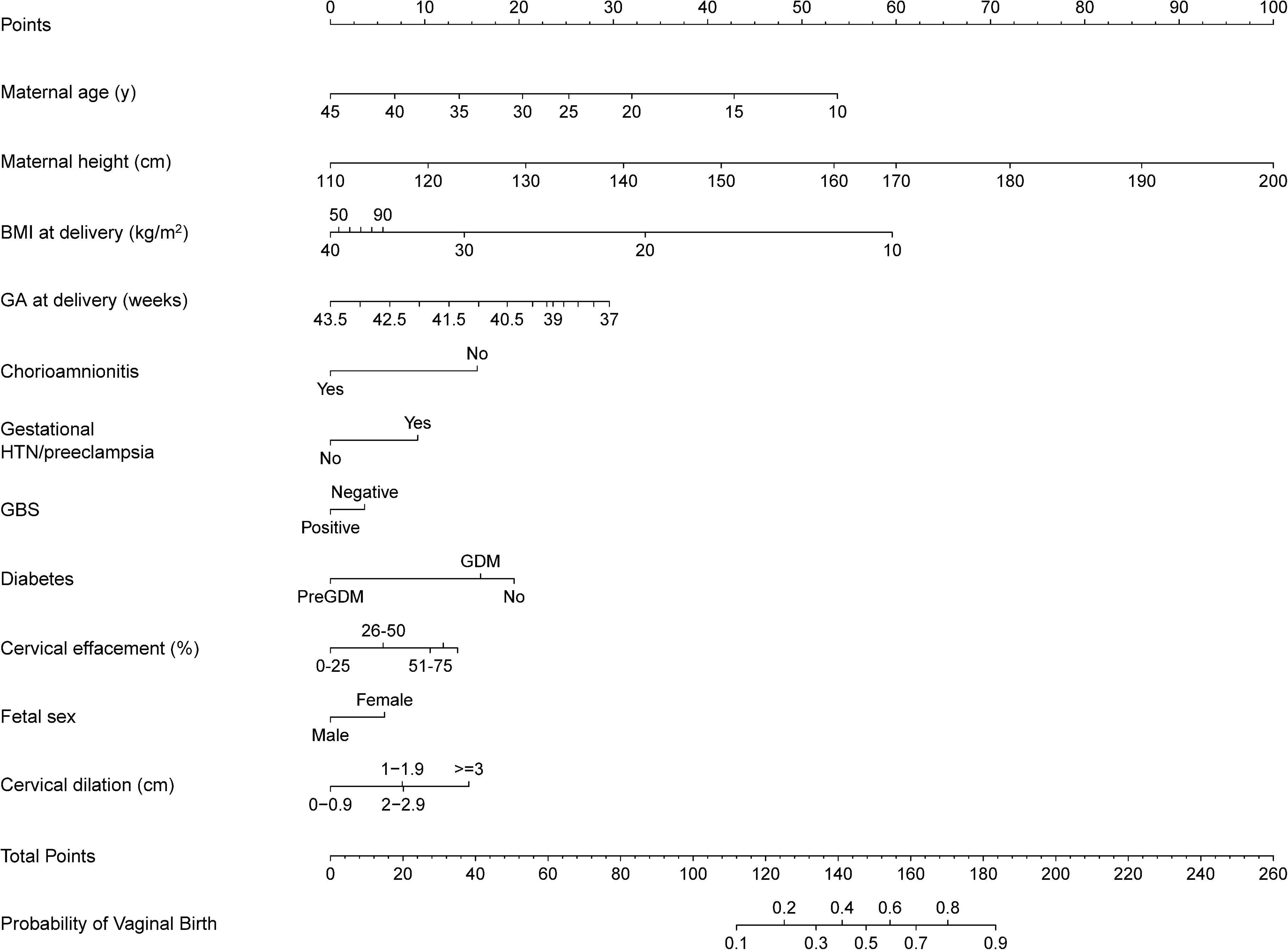
Nomogram to predict the probability of vaginal birth in term nulliparas. To use this nomogram, locate the patient’s characteristics and their corresponding number of points using the “points” scale. Sum these points identified from each of the eleven predictors, and locate the sum on the “total points” scale. The sum on the “total points” scale corresponds to the probability of vaginal birth in term nulliparas provided on the “probability of vaginal birth” scale. Abbreviations: BMI, body mass index; GA, gestational age; HTN, hypertension; GBS, group B streptococcus; PreGDM, pregestational diabetes; GDM, gestational diabetes.

## Discussion

This study aimed to detect factors that can accurately predict successful vaginal birth in nulliparous individuals at term. Independent predictors of vaginal birth were identified and included maternal age, maternal height, BMI at delivery, gestational age at delivery, GBS status, chorioamnionitis, gestational hypertension/preeclampsia, diabetes, cervical dilation, cervical effacement, and fetal sex. When these factors were all incorporated into a regression model, the resulting ROC curve failed to accurately predict vaginal birth as the AUC was <0.80.^34^

Previous attempts at developing prediction models for route of delivery resulted in moderate predictive capacities with AUCs ranging from 0.709 to 0.790.^15, 27–29, 31^ Consistent with our study, these models, most of which also explored nulliparous populations, found that younger maternal age,^14, 15, 28, 31^ greater maternal height,^28^ lower BMI at delivery,^27, 31, 35^ earlier gestational age at delivery,^27, 28, 31^ the absence of pregestational diabetes,^28^ greater cervical dilation,^28, 31^ and greater cervical effacement^28^ were associated with vaginal birth. Likewise, studies examining predictors of cesarean delivery reported associations between cesarean delivery and older maternal age,^14, 15^ shorter maternal height,^15, 29^ greater BMI at delivery,^15, 29^ later gestational age at delivery,^15^ pregestational diabetes,^15^ hypertensive disorders,^15^ lower cervical dilation,^15^ and male fetal sex.^15, 29, 36^

Contrary to previous findings, the present study did not find associations between vaginal birth and race,^28^ health insurance,^14^ gestational diabetes^15, 28^ and chronic hypertension^28^ after adjusting for possible confounders. Discrepancies in findings may be attributed to variations in patient selection and outcome assessment as these previous models were designed to predict vaginal birth in specific patient populations. For instance, Kamlungkuea *et al*.,^27^ Kawakita *et al*.,^28^ and Tolcher *et al*.^15^ included patients with induction of labor only, excluding spontaneous labor.^37, 38^ We included both spontaneous and induced labor since spontaneous labor comprised a majority (60.9%) of the vaginal births in our cohort. Similar to Costantine *et al.*’s findings,^31^ our study revealed that induced versus spontaneous labor did not play a role in predicting vaginal birth. Kamlungkuea *et al*.’s predictive model of vaginal birth in Thai patients included not only induction of labor but also parity.^27^ Multiparous individuals with a history of vaginal birth have been shown to be more likely to successfully deliver vaginally,^39^ which will inevitably impact the prediction model. Costantine *et al*. limited their analysis to patients without any obstetric or medical comorbidities, thereby limiting the model’s applicability to high-risk nulliparas that are at risk of cesarean delivery as shown previously. ^15, 28^ Our analysis addressed the limitations of Kamlungkuea *et al*. and Costantine *et al*. by including only nulliparous individuals and individuals with comorbidities.

Regarding our finding of positive GBS status decreasing the odds of vaginal birth, to our knowledge, only one study evaluated GBS status as a possible predictor of cesarean delivery, and an independent association was not detected.^15^ Our findings may be explained by the increased risk of chorioamnionitis associated with GBS colonization and a 1.39 fold increased risk of cesarean delivery associated with chorioamnionitis.^40–42^ Our study showed that chorioamnionitis was associated with a reduced probability of vaginal birth, and a significant correlation between positive GBS status and chorioamnionitis was also detected in the study sample (*p*=.003).

### Clinical and Research Implications

Our model was designed to assist and supplement, not replace, the process of decision-making regarding the route of delivery by providing patient-specific risk assessments to both the clinician and the patient given nulliparas’ lack of prior obstetric history. It was developed to guide labor management at the time of admission, particularly for patients at increased risk for cesarean delivery. For example, a clinician can still offer a trial of labor to a patient with a low probability of vaginal birth based on our model. However, the clinician would know beforehand not to prolong induction or the second stage of labor beyond reasonable recommendations.

Our model included one modifiable factor: BMI at delivery. Hypertensive disorders of pregnancy and pregestational diabetes were also identified as independent predictors. Thus, one can recommend lifestyle modifications, healthy diet, smoking cessation, and physical activity, to prevent excessive gestational weight gain, diabetes, and cardiovascular disease, and hence increase the odds of vaginal birth.^43, 44^ Future research directions include external validation of the model before implementation in clinical practice, after which subsequent pregnancy outcomes can potentially be investigated.

### Strengths and Limitations

Strengths of this study include the large and racially, ethnically, geographically diverse group of nulliparous pregnant patients analyzed. The distribution of race-ethnicity of our cohort is comparable to that of the general population in the United States in 2023, which improves the generalizability of our findings.^45^ The mean age of first-time mothers in 2021 was 27.3 years,^2^ which is also comparable to the mean age of our cohort. In addition, our model included factors not only readily accessible to the obstetric provider but also clearly defined and routinely determined at the time of the delivery admission to increase its clinical utility. Our predictive model can be used to provide nulliparas seeking vaginal birth with the probability of its success using factors usually present at the time of admission, and it can encourage obstetric providers to aim for vaginal delivery in the absence of a medical indication for cesarean delivery. Finally, our model generated an AUC greater than most previous models with high precision given the narrow 95% CI.

Limitations of the study include the lack of clinical utility of the final model. Although internally validated, the model must be validated in different patient cohorts and settings and compared with other models. The potential effect of residual confounders that have previously been shown to affect mode of delivery, such as attitudes towards mode of delivery and physician cesarean delivery rates, cannot be disregarded. However, any factors that cannot be measured objectively and/or accurately documented were purposely excluded during model development.

## Conclusion

Several factors available prior to labor were identified to be independently associated with vaginal birth among term nulliparous patients. The overall final model performed with fair predictive capacity but cannot be used to accurately predict vaginal birth in term nulliparous individuals at the time of admission. Optimizing the success of vaginal births in nulliparas and promoting their overall obstetric care to reduce cesarean delivery rates still requires additional research.

## Data Availability

The data is from the Nulliparous Pregnancy Outcomes Study: Monitoring Mothers-to-be that is hosted on https://dash.nichd.nih.gov/study/226675

